# An exploration into the impact of the COVID-19 pandemic on maternal mental health in high-and middle-income countries with a case study in East Sussex

**DOI:** 10.1101/2023.11.02.23298002

**Authors:** Abbeishna Sabesan, Caroline Ackley, Sophia Stone

## Abstract

**Background:** The physical isolation that the Coronavirus pandemic enforced resulted in a decline in mental health that disproportionally affected higher risk individuals, including women in the perinatal period. The wellbeing of perinatal women was, at times, neglected due to hospital and governmental isolation regulations. The aim of this study was to conduct a scoping review and Key Informant Interviews (KII) to identify gaps and opportunities for further research, and to distinguish discrepancies and correlations between the two sources of information.

**Methods:** Two methods were utilised: a scoping review and KIIs. The scoping review identified relevant articles through a database search on Google Scholar, PubMed and EBSCO. The KIIs were conducted virtually with counsellors working in a maternal mental health charity in South-East England. Both methods collected qualitative data and were thematically analysed.

**Results:** 95 articles were eligible for the review and 5 participants were recruited for the KIIs. Thematic analysis revealed 6 themes in both sources (1) demographics; (2) support; (3) policy; (4) insecurity; (5) anxiety; and (6) milestones. Between the two datasets there were no disparities in the impacts of changing policies, fear of the virus, and the grief caused by missing perinatal milestones on mental health. Significant discrepancies were identified in the influence of demographic characteristics, which was a risk factor for adverse mental health outcomes.

**Conclusion:** The most prominent theme in the review is the reduced support available, whilst the KIIs indicate that changing hospital policies are the main cause of harm to perinatal mental health. Birth trauma is deemed to be of significance in the interviews but not in the literature. Further research should focus on the impact of the pandemic on recovery following birth trauma to identify long-term implications and facilitate policy changes to reduce the risk of birth trauma/ post-traumatic stress disorder.

## Introduction

The Coronavirus (COVID-19) was first reported in December 2019 and quickly spread to become a global pandemic resulting in unprecedented times. Physical isolation by the public was required to prevent further spread of the virus (1). The enforced isolation had a significant effect on mental well-being, particularly impacting high risk populations. Research by Hermann et all (2021) highlighted that perinatal women, encompassing those who are pregnant, giving birth, or in the first year of parenthood, are among those most affected (2). They are therefore more vulnerable to mental health illness. During the postpartum period, women experience a multitude of life transitions, examples including a transformation in personal identity, and major changes in the dynamics of the relationships with their partner, family, and social life in general. These changes can contribute to negative psychological states and the development of depression, anxiety, and post-traumatic stress disorder (PTSD) (3).

The precarious situation new mothers find themselves in was exacerbated by the additional pressures of the pandemic. The pandemic left the UK government with no alternative but to enforce physical distancing and isolation. Hospitals implemented essential policies to manage the steep influx of COVID-19 patients and to prevent the spreading of the virus; this had a negative psychological impact on patients (4). Policies were implemented in all hospital departments including the maternity and delivery wards where mothers gave birth in the absence of their partners, experienced reduced postnatal hospital stays, and had limited postnatal visitation hours. The combination of government and hospital policies deprived parents of their experiences and expectations of birth and postnatal plans they originally anticipated. These lost experiences may have been as simple as having their baby bump acknowledged, or having family and friends visit their baby at the hospital, and even as fundamental as having the assistance from healthcare professionals (5).

Positive maternal mental health is paramount for both mother and baby and can be impacted by adverse pregnancy events with consequent social implications for the baby. Compromised mental health has been linked to obstetric outcomes including, preterm births, fetal growth impairments, pre-eclampsia, and antepartum and postpartum haemorrhages has been explored (6). Furthermore, the long-term consequences on children include requiring tailored educational needs, lack of formal qualifications, and development of emotional and behavioural conduct problems (7).

A systematic review in 2015 assessed the socio-economic implications of maternal mental health disorders in the UK and showed that the total lifetime cost per woman with perinatal depression and anxiety was £75,728 and £34,811 respectively, which accounted for the cost of health and social care, education, informal caregivers, and criminal justice, as well as losses in quality of life and productivity, for both mother and child. This totalled a countrywide expenditure of £6.6 billion, of which 60% was owed due to the economic burden associated with childhood morbidity (7).

### Aim and objectives

This study aims to explore the impact of the COVID-19 pandemic on maternal mental health in the UK and HMICs, with a particular focus on South-East England. It intends to identify areas in the field of maternal mental health that have been under-researched and overlooked. To do this, a scoping review was conducted along with Key Informant Interviews (KII) with counsellors who provide mental health support to new mums in the South of England. Gaps and opportunities for further research were identified as well as, discrepancies and correlations between the two databases.

### Rationale

A preliminary search on PubMed and Google Scholar was conducted prior to the research study and it was found that there was no scoping review data on maternal mental health from pregnancy to 1-year postpartum during the COVID-19 pandemic. The search also showed predominantly quantitative research with only speculation into causes of poor maternal mental health. In response, this research focused on the analysis of qualitative data to ascertain a description of experiences faced by pregnant women and new mothers during the pandemic. Participants in the studies included in the scoping review were predominantly middle-class, Caucasian, heterosexual women; minority groups are grossly unaccounted for in published data. By conducting the KIIs in the South-East of England, we can investigate a similar demographic and therefore detect any discrepancies present in the studies outlined in the scoping review. Moreover, no studies considered the views and insights of counsellors who supported new mothers during the pandemic and the impact this had on maternal mental health outcomes and experiences. Their experiences are important because counsellors and mental health professionals have first-hand knowledge on the wellbeing of new mothers and can provide recommendations to improve maternal mental health outcomes. Finally, many women who had a baby during the pandemic may be pregnant or planning for a subsequent pregnancy and interviewing counsellors will allow for insight into differences in contributors of maternal mental health pre-pandemic, during the pandemic, and post-pandemic.

## Methods

This study utilises two main methods: (1) scoping review and (2) KIIs. The scoping review was conducted to gain an understanding of the current research on maternal mental health during the pandemic and to identify research gaps in existing literature. KIIs were conducted with counsellors located in South-East England working in a women’s mental health charity during the pandemic. The interviews were conducted in February 2023.

### Ethics

Ethical approval was not required for the scoping review component of the research but was needed for the KIIs. Ethical approval was granted for the KIIs by the Brighton and Sussex Medical School (BSMS) Research Governance and Ethics Committee (RGEC) on 16 January 2023 (ER/BSMS9EKA/1).

### Scoping review

This review followed the methodological framework of Arksey and O’Malley, consisting of five main steps: identifying the research questions, identifying relevant studies, study selection, charting data and finally collating, summarising, and reporting the results (8).

### Step 1: Identifying the research question

The research question that guided the scoping review is, *what is known about the impact of the COVID-19 pandemic on maternal mental health in the UK and HMICs?* The study population included pregnant women and mothers up to 12 months postpartum. The intervention investigated was the COVID-19 pandemic and this was compared to the situation pre-pandemic. The outcomes explored are the psychological and emotional effects of the COVID-19 pandemic on pregnant individuals and new mothers.

### Step 2: Identifying relevant studies

A literature search of studies published between 2019 and 2022 was conducted using Google Scholar, PubMed and EBSCO. Table 1 shows the search query used in the databases.

**Table 1:**
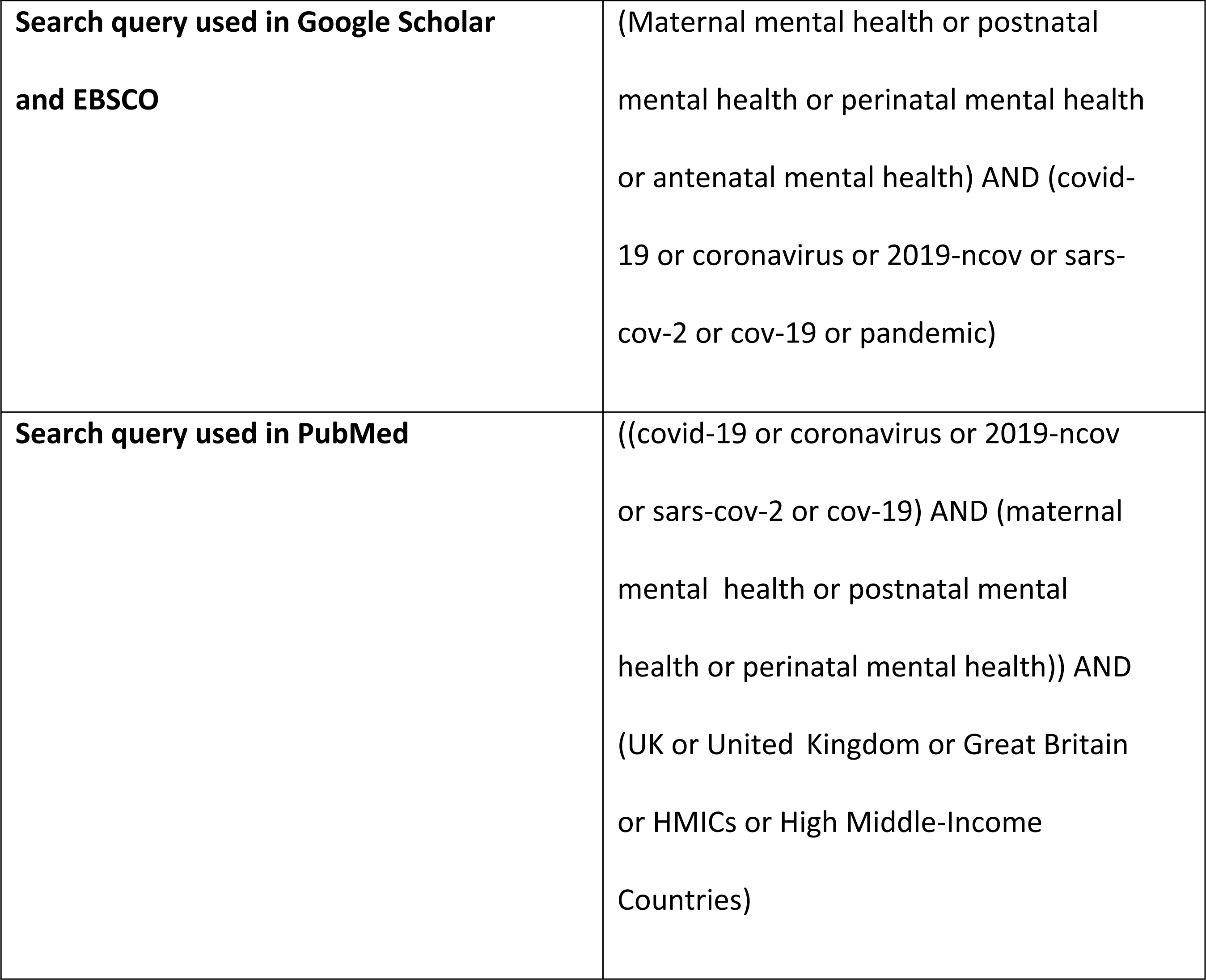
Search query used in the databases.

### Step 3: Study selection

The studies were screened in a 2-stage process. First, studies were screened based on their title and abstract to decipher whether it fit the inclusion criteria, which consisted of: (a) population of pregnancy to first 12 months postpartum, (b) studies in HMICs and (c) in English. Editorials, letters, commentaries, protocols, and articles without full-text coverage were not eligible for inclusion. Then, the following questions were asked for each study:

1. Is the article related to maternal mental health in the time of the COVID-19 pandemic?
2. Is the article related to a factor affecting maternal mental health?
3. Is the article about restrictions put in place at the time of the pandemic?
4. Is the article about changes in the care of pregnant women in the time of COVID?
5. Is the article about the care of women in the postnatal phase?

The article must have met the inclusion criteria and agreed with at least one of the above questions. Articles not meeting this requirement were not considered any further, whereas eligible studies were classified in a database.

The second screening stage included using the CASP qualitative checklist to assess validity, relevance, and significance of potential articles. A score of 7 or greater was needed to pass the checklist and to be included in the study, whilst those scoring 6 or lower were excluded. CASP scores were included in the database.

### Steps 4 and 5: Data collection and collating, summarising, and reporting results

Data was extracted from eligible studies into the database to summarize the following: title, author, year of publication, study location, aims, methodology, outcomes, and CASP score. Finally, the remaining articles were analysed using thematic analysis. This analytical approach was used because of the time-efficiency it provided and the flexibility to explore a wide range of concepts (9). Themes were identified according to their potential impact on maternal mental health, which was then included in the database.

### Key Informant Interviews

KIIs were conducted with five participants to gain a better understanding of maternal mental health during the pandemic in South-East England and to provide a comparison to the scoping review results. KIIs were carried out in the South-East of England because this was the base of the investigators conducting the study and recommendations developed from the study could be shared with local stakeholders. A semi-structured interview guide was developed based on themes that emerged from results of the scoping review. The interview guide consisted of open-ended questions that followed a flexible format, thus allowing relevant issues to be discussed in-depth and for new themes to emerge.

Participants work in a mental health charity that provided support to new mothers during the pandemic and were recruited through a gatekeeper. The charity delivers 10-week postnatal therapeutic groups to mothers up to one-year post-partum. The locality of this organisation made this a practical choice in its cooperation in the study. Approval was gained and potential interviewees were obtained following recruitment email and participant information by the gatekeeper thus ensuring the research team were not involved with participant selection. The recruitment period was between 17/01/2023-6/02/2023. The inclusion criteria for the participants were (a) age >18, (b) consented individuals (c) worked and working in the field of maternal mental health during the COVID-19 pandemic. Five participants responded and were eligible to participate in the study.

Prior to each interview, verbal consent was obtained, and was documented at the beginning of their transcripts. The KIIs took place over Microsoft Teams and the duration ranged from 45 to 60 minutes and were all audio recorded and transcribed by the lead researcher.

Transcriptions were read, reviewed, and qualitatively analysed using thematic analysis. Key quotes and its analysis were entered into a database, where its significance was defined, along with a comparison to scoping review data.

## Results

### Thematic analysis

Six themes were identified during analysis of the literature included in the scoping review and the KIIs: (1) support; (2) anxiety; (3) insecurity; (4) policy; (5) milestones; and (6) demographics. Convergence was seen between all the themes. The theme, ‘support’ was further split into 4 subthemes: (1) social; (2) support groups; (3) domestic support; and (4) healthcare. The issues surrounding healthcare support will be discussed in the ‘policy’ section as much of the results intersected. Data collected in both methods for each of the above themes will be described separately.

### Scoping review results

#### Search results

As represented by the flow diagram in Figure 1, the systematic search identified 687 studies. The initial relevance screening excluded 548 studies and a further 34 were removed due to duplication, leaving 105 studies to enter phase 2 of eligibility screening. 10 studies were excluded at this stage because they did not score 7 or more with the CASP qualitative checklist, leaving 95 studies to be included in the scoping review.

**Figure 1:**
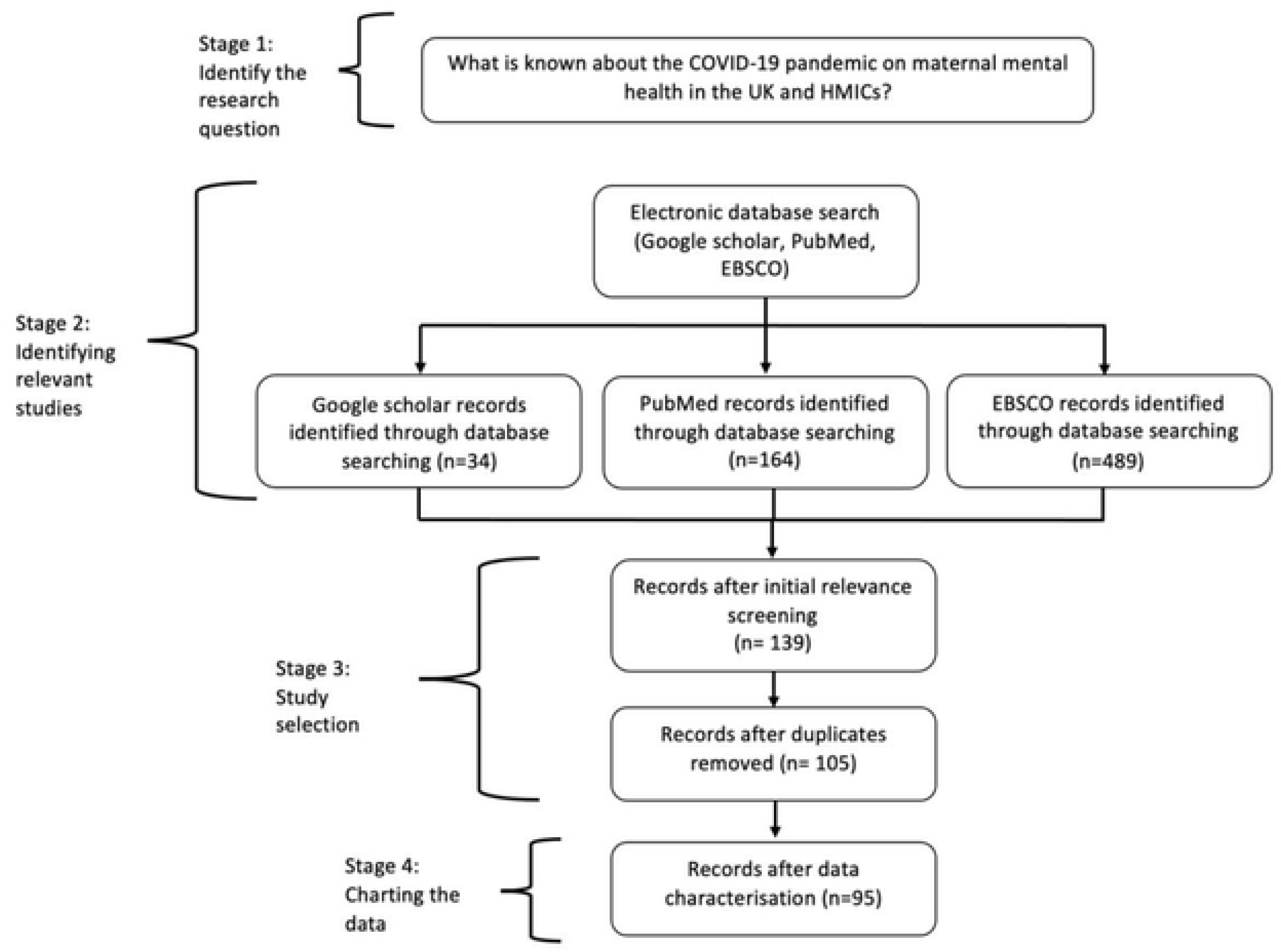
PRISMA flow diagram of studies identified, screened, and extracted

#### Demographics

Education level, ethnicity, age, parity, pregnancy stage, employment, income, and socioeconomic status are a few of the demographic characteristics investigated. Some studies highlight different characteristics, like income to be significant (10), whilst others claim that demographics, as one entity, played no role in maternal mental health (11). Three studies state that having a higher education level results in less anxiety and depressive symptoms, and a more supportive experience perinatally (12–14). However, one study claims that there is more anxiety and stress in this group of people (5). Moreover, all studies agree that women of colour are overall disadvantaged compared to their Caucasian counterparts; they feel less supported, find healthcare to be less accessible, they are more worried about employment and are more depressed and anxious (12, 16–19). Other demographic characteristics associated with poorer outcomes are (1) having a lower socioeconomic status, (2) previous mental health diagnosis, (3) having a low income and (4) being unemployed (17, 20, 21). Studies have differing results regarding the impact that age, parity, and pregnancy stage have on maternal mental health.

#### Support

Fifty-one studies investigate the role of ‘support’ on perinatal mental health; below is the data collected for the subthemes listed above (1, 5, 10, 12, 14, 17–19, 22–60).

#### Social support

Support from family and friends lacked considerably throughout the pandemic; a total of thirty-nine studies highlight the importance of this (5, 10, 12, 14, 17–19, 23, 24, 26–33, 35–39, 42–44, 46,54, 56, 57, 59–61). Social support is proven to be protective against poor mental health outcomes both in pregnancy and postnatally (36, 51, 53). One UK interview-based study notes the sleep deprivation that follows a lack of support, “I think week two to four was peak tiredness and then that’s the point where I’d really loved either my mum, my mother-in-law, or my own family to sort of step in and be able to help out a bit more” (29). The same study notes the frustrations some women expressed, “particularly with my mum, she’s been self-isolating. We’ve been self-isolating…I can’t see why we can’t see family” (29). However, three studies note that some women appreciate the time they spent without their social network (5, 12, 49), “Women reported having a new appreciation for quiet moments, family time, and a slower pace. Some expressed appreciation for stay at-home requirements that allowed more time together with their partner and more sharing of parenting responsibilities” (5).

#### Support groups

Ten studies mentioned the importance of attending parenting groups and their potential benefits, such as meeting and forming friendships with new mothers (17, 18, 25, 29, 34, 39, 51, 52, 58, 61). Parenting groups are seen as a place to get invaluable social exposure and advice associated with improving one’s emotional wellbeing (29). Although, during the pandemic some groups ran online, most women found that the in-person experience was irreplaceable (25).

### Domestic support

Issues surrounding domestic support were discussed in thirteen studies (14, 17, 19, 23, 25, 27, 31, 37, 38, 41, 46, 51, 52). Data shows that women tend to bear a larger burden than men in addressing these pressures and was proven in a Canadian study that fifty more hours were spent by the average woman on childcare during the pandemic as opposed to the average man (14). The bearing of this workload was established in a US nationwide study where it was found that 56.3% of women found the lack of childcare to be stressful (41). Managing the demands of their newborn whilst also supervising their older children’s home-schooling was taxing. Guilt was often felt as time spent with older children was compromised compared to that of younger children (25).

#### Policy

The issues surrounding the implementation of hospital and governmental guidelines was raised in twenty-two studies (1, 4, 12, 23, 24, 26, 29, 32, 34, 37–40, 44, 45, 53, 55, 62–65). Policies were everchanging and not well communicated by hospitals, causing confusion and uncertainty; birth plans were altered to satisfy these new regulations (4). Several policies did not permit visitors following the birth, and the presence of the partner was disallowed in appointments and at the birth itself (38). Additionally, seventeen studies discuss the issues surrounding an absent partner (17, 19, 24, 25, 28, 30, 32, 34, 37, 38, 40, 44, 52, 54, 55, 59, 60). For example, a study including 6894 participants in 64 different countries, shows that these hospital policies, affected 41% to 59% of mothers (32). Maternal healthcare was grossly inaccessible and appointments were constantly being cancelled (55). The appointments were primarily over the phone and often rushed, triggering a sense of abandonment and dismissal in pregnant women (24). Face-to-face appointments were uncommon, but when they occurred, healthcare professionals were in PPE and were told to avoid having any physical contact (34). Overall, the literature suggests that mothers felt unsupported by healthcare professionals and a total of twenty-one studies alluded to this (1, 12, 17, 18, 23–26, 28, 29, 32, 34, 37–39, 41, 44, 45, 55, 57, 58).

### Insecurity

Insecurity, encompassing feelings of uncertainty, was common and described in twenty-one studies (4, 5, 13, 18, 21, 23, 25, 26, 31, 32, 36–38, 41, 66–70). Job and financial security were a major source of concern in the pandemic, with women mainly worrying about their partner losing their job and not being able to provide for their baby (4, 13, 25). For some, this unemployment resulted in the inability to buy clothes for their child and instead rely on presents they had been given (68). The UK’s furlough scheme, a policy in which employers received 80% of their salary from the UK government and temporarily stopped employee wages, was mentioned in one study and stated that it mitigated this situation in the latter parts of the pandemic (71). Financial security also includes women either losing their own job or being forced to reduce their working hours due to unproductivity at home that followed managing new responsibilities of this new working environment, such as childcare (37). Moreover, a limited availability of essentials such as food, medicines, and baby supplies, for example nappies and formula, was also a source of insecurity. A US study found that of the 2740 surveyed pregnant women, 59.2% and 63.7% were stressed about food running out and losing their job or household income respectively (41).

#### Anxiety

Thirty-five studies investigated perinatal anxiety which was often in reference to a mother’s fear of the virus, leading them to follow the news and social media excessively to gain clarity of the situation, which in turn led to further enhancement of this anxiety (1, 4, 5, 18, 19, 23, 25, 26, 28–32, 35–37, 39–41, 43, 44, 52, 54, 56, 58–60, 68, 72–77). One US study proved that time spent watching COVID-related media content was linked to mental health symptom severity. Those viewing this content for more hours in a day were more likely to experience anxiety symptoms (5).

The literature states that dilemmas were faced at times when certain worrisome pregnancy symptoms were experienced, which would normally lead them to go to the hospital or maternity assessment unit, however mothers found themselves too afraid of going (29). Women most feared that either themselves, their baby, or loved ones would contract COVID; this was of concern not only during the pandemic, but also in the period of which there was easing of restrictions and social distancing (25).

With reduced support from family and friends, there was anxiety around the competency of their own parenting, as well as the wellbeing of their baby and its impact on their development caused by the lack of exposure to other children and people (35). Anxiety was also the result of certain insecurities mentioned previously namely, scarcities in food and uncertainties in employment (23).

#### Milestones

The grief experienced by parents because of missing key milestones was articulated in eight studies (5, 19, 29, 31, 34, 64, 68, 78). Antenatally, women were forced to cancel or have virtual baby showers, as well as miss out on attending in-person birthing classes (19, 34). They were deprived of the opportunity to shop for baby furniture and clothes and were unable to display their baby bump (34). Furthermore, some felt that their maternity leave had been wasted as everyone was staying at home already (31). Postnatally, their baby was unable to form relationships with grandparents or other loved ones (29).

Ultimately parents felt as if they had missed out on celebrating the birth of their child and received an inadequate new-born’s experience (64).

### Key Informant Interview results

#### Participant characteristics

Nine counsellors were contacted by the gatekeeper, seven of which responded and five met the inclusion criteria and were interviewed by the lead researcher. All the participants were trained counsellors with various roles in the charity, including two lead facilitators, counsellors who run group therapy sessions; two support facilitators, who run smaller group discussions; and the founder. Their time spent working in the field of maternal mental health ranged from two to over ten years.

#### Demographics

Four participants concluded that a higher education level resulted in better maternal mental health outcomes. For example, one participant considered mothers in town A to be better educated and of a higher socioeconomic group than those in town B, “The mums in town A knew how to stay calm, and they knew more about their babies, and perhaps read more. But mums in town B, were more hopeless, when it came to things like that. They didn’t have the knowledge to inform what they were going through” (Participant 2). The same participant recounted her experience at a distressing support group she led in town B, “I noticed more in town B that the voices of the mums were less heard. I don’t know if this was because they didn’t feel able to ask for what they needed, maybe less sense of self-importance, so maybe they didn’t challenge some of the things the hospital was saying versus the people in town A whose self-worth was higher. In one group at town B, every mum was crying. This was the most traumatic group I’ve ever done.”

Four participants showed discomfort when questioned on the impact of demographics on maternal mental health, “I find these questions difficult because I feel bad, it’s patronizing. I find this difficult to talk about” (Participant 5).

#### Social support

A lack of social support was expressed by four interviewees, “Suddenly mums became very isolated from support networks who would have normally helped a new mum practically and emotionally. All the pressure was on the parents to do everything” (Participant 4). This ultimately resulted in mothers compromising their selfcare, “Mums say managing to have a shower is a huge achievement. Some are able to do this if they have supportive partner” (Participant 1). Participant 5 states that lack of social support was not mentioned as “They didn’t know any different”. She went on to say, “The extra family support is really important in the early days, but they didn’t realise that. So, they’re hard on themselves and think they’re doing a bad job, but they don’t realise how challenging it is to do by themselves”.

#### Support groups

KIIs stressed the value of attending support groups as attendees were assured that their experience was being reciprocated by mothers in a similar same situation, “it was refreshing for their feelings to be normalised and to hear other mothers were in line with what they were feeling” (Participant 2). Furthermore, attending support groups “improves their confidence and their ability to regulate their anxiety and low mood” (Participant 1). Another KII reveals that support groups are potentially more valuable than the support from family and friends, “When people share birth experience with friends and family, they put a positive spin to it, and make mums feel like they can’t share, but being in this space will allow people to hold that grief” (Participant 1). On the contrary, one participant revealed a negative side of support groups, “It’s competitive or they just put on a brave face as they want people to think that they have it all together through social media” (Participant 1).

#### Domestic support

The issue of increased domestic work was brought up by two participants, “During the pandemic, I knew mums who needed to keep a house in order, home-school other children, manage employment. All these added pressures, whilst not being able to receive support to refuel themselves…Not being able to go fully outside to enjoy things that can be replenishing, for example exercise being a taken away, added to the intensity to the home” (Participant 4). This increased workload led to, “Feelings of shame and guilt that they couldn’t keep up with it. It’s harder to leave their space, so you’re sticking with the mess” (Participant 5). On the contrary, two participants stated, “The pressure was left a little bit. No one’s going to come over. It (domestic work) didn’t make a difference or to a certain extent there was less pressure” (Participant 4).

#### Policy

All participants discuss the effects of changing policies. Disallowing partners at the birth and at antenatal appointments meant that, in the case of receiving bad news, such as the loss of a pregnancy, women were unaccompanied. Also, birthing alone required them to advocate for themselves at a time of vulnerability: “They’re frightened, they don’t know if this is normal. They’re in pain, wondering what their option for pain relief is with no doctor or midwife there in that moment” (Participant 4). One support facilitator recounts a traumatic birth story of one mother; the quotation demonstrates the effect of an absent support partner, who would have made logical decisions in the best interests of the mother, “A young girl had been given a really strong painkiller, against her will.

Against their will, in the sense that when you’re in labour and you feel that you don’t have choice, partner wasn’t there, and they made it seem like you had to take this. The way she saw it was like they were sedating women because they were so overcrowded that there was nothing they could do. She really didn’t want to take it, she said it made her feel really ill, but she also couldn’t remember the first few hours of her baby being born because it was such a strong drug. A few other people have said similar things where they were given strong drugs which they wouldn’t normally accept, and they wouldn’t normally have been rushed to take them but did because of the overcrowded element” (Participant 5).

Postnatally, mothers were not allowed visitors, leaving hospital staff to assist them more than they typically would, “Normally, family would help the mum, bring food and going to the toilet. All of that was on the staff but they were just so inundated with so much to do.

So that was a policy, they ended up changing to just one person being allowed to visit for limited periods of time. All of these polices really impacted people feeling supported at a really vulnerable time having just given birth” (Participant 4).

Interviewees described the situation of lacking hospital support before the above policy modification, “They were left at a scary time when they’ve just had a baby and you’re hurting physically and no one’s there to get a glass of water or pass the baby and you have to do it all yourself and your partner can’t be there” (Participant 2).

Additionally, moving from a homebirth to a hospital birth was a change that often occurred. One KII participant explains, “You’re trying to avoid the medical environment. It was bad pre-pandemic, but then in the pandemic, with all the tests and wearing masks, if you’re already inclined to not have this part of your birth, it must have been harder in the pandemic because it was especially medicalised” (Participant 5).

Rushed and inaccessible healthcare further exacerbated the usual anxiety that comes with becoming a new mother, “Concerns weren’t really addressed. They had no one to turn to for help. When you’re a new mum you’re so anxious about everything, like am I feeding enough?” (Participant 2). Additionally, the midwives wearing PPE resulted in, “not having the tactile support that sometimes mums need. Everything was distanced and removed a sense of support” (Participant 4).

#### Insecurity

One participant speaks about financial concerns, “One mum in particular, whose partner was working in advertising and because events stopped, she made the decision of going back to work much sooner because her career was more stable. So, it created a lot of distress, especially for mums who weren’t ready to go back” (Participant 4). The interviewee, with over ten years of experience, explains that often maternity leave acts as a reflection period during which mothers can question their professional identity.

Therefore, shortening maternity leave, restrains mothers in making potentially key life decisions. Participants 2 and 3 state that financial security is insignificant, “I think if people were furloughed, it wasn’t a terrible thing. No, I don’t think this was much of a worry” (Participant 3). Moreover, two participants expressed mothers worrying about the insufficiency of resources, “They felt insecure about getting food, toilet roll, nappies” (Participant 3). Feelings of insecurity and unsafeness at home was raised, “A couple of mums at some point during the pandemic, said their relationship had become abusive” (Participant 2).

#### Anxiety

All participants discuss the anxiety that came with contracting COVID, “Would my baby be okay? Would I be okay? … They were very afraid of taking their child out of the house. This had a knock-on impact as this meant that mums were being stuck at home… but also staying at home made them anxious because it fed into them feeling isolated” (Participant 4). KIIs indicate that there was anxiety related to rules being ambiguous and even though mothers didn’t feel comfortable socialising, they were unsure on how to deal with those who wanted to. The result of this anxiety was conveyed in the interview of a lead facilitator working in the field for over three years, “A lot of people were having dark intrusive thoughts, meaning they were constantly thinking something was going to happen to them or their baby. Mums said that they used to be walking down the streets, and they thought that a car was going to crush them. This doesn’t happen anymore, but certainly heighted in the pandemic.” (Participant 2).

#### Milestones

Grief surrounding missed milestones was universal to all five participants, “No one saw them pregnant, no one celebrated their pregnancy. They didn’t feel that they could show off their bump. Couldn’t have baby showers. Some people didn’t meet baby until two or three months and even then, they were scared they would give COVID to baby” (Participant 2). Another interviewee took this further by stating that, “This idealized time of becoming parent wasn’t what they wanted. So, normalizing that this was a loss, and feelings of anger is normal and makes sense …There was a real sense of sadness that came out with symptoms such as depression” (Participant 4).

### Strengths and Limitations

Our recruitment strategy minimised chances of selection bias as a gatekeeper was appointed to select potential interview participants. Bias arising from individual studies included in the scoping review was minimal owing to the use of the CASP qualitative checklist. Nevertheless, the study has limitations that must be addressed. Interviewees were expected to recount past encounters, introducing recall bias which may have led to either the over or under exaggeration of data (80). Inclusion bias may have been introduced since only articles published in English were included, potentially reducing the breadth of data analysed. The internal validity may have been compromised as there was a single investigator selecting studies and analysing data, rendering the study susceptible to selection bias (81). Additionally, the interview guide was developed based on the themes extracted from the scoping review. Whilst this ensured that the interview explored relevant concepts, this could have potentially limited the emergence of new themes and constrained the findings to what had already been explored. Moreover, time constraints limited the number of organisations approached and consequently the number of participants included in the study. Recruitment of more experts would have enhanced the validity of the findings.

## Discussion

Several key themes were identified in the KIIs and the literature review, however the most prominent theme in the literature review was the reduced support available to mothers, (57% of the studies). Whereas KIIs indicate that changing hospital policies was the most detrimental to perinatal mental health. This divergence emphasises the complexity of perinatal mental health and demonstrates that no single theme can fully encapsulate the spectrum of influences on mental health during the perinatal period.

The influence of various demographic characteristics on maternal mental health was inconclusive; being of an ethnic minority was the only characteristic deemed significant in causing poorer mental health outcomes. Evidence from the KIIs was limited as participants predominantly worked during the pandemic with first-time mothers of a British, Caucasian, and middle-class background, providing no substantial demographic diversity.

There is therefore merit for further investigation which will help to identify demographic characteristics that render a woman vulnerable to poor mental health outcomes. This could facilitate the establishment of an antenatal screening programme aimed at identifying individuals at risk, in doing so provide early support during their pregnancy, to prevent any long-term sequalae of delayed identification of mental health conditions.

Moreover, amongst the interviewees, there was an underlying discomfort in discussions surrounding demographics, which implies that there might be an underlying stigmatisation which they find difficult to portray; further investigation needs to go into this.

KIIs and literature data reveal that poor birth experiences were caused by rules inhibiting a birth partner, as well as the inaccessibility and intangibility of healthcare, which ultimately resulted in insufficient care and a distrust in medical professionals. Additionally, there is substantial evidence in both databases that indicate that the obscurity of the virus caused fear and, in some cases, a paranoia about unlikely incidents. A longitudinal study exploring the long-term psychological consequences of the pandemic, would be a valuable addition to the database, for example whether COVID regulations resulted in the development of new behaviours such as a health seeking avoidance.

While both the literature review and the KIIs stress the negative implications of hospital policies, only the KIIs discuss birth trauma to be a consequence of it. Interestingly, all five participants observed the pronounced influence the pandemic had on birth trauma, while the literature review did not. Therefore, this provides new insight into the relationship between policies and mental health. Investigation into the impact of the pandemic on birth trauma should be coordinated to identify any long-term implications. This research will provide a solid rationale for any necessary policy changes adopted in the future to reduce the risk of birth trauma and consequently, mitigating subsequent repercussions.

Bethany Kotlar et al demonstrated that the incidences of stillbirth and neonatal mortality were significantly higher during the pandemic (23). Although multifactorial, contributing to this higher incidence during the pandemic, was the reduction in antenatal and postnatal appointments and the reluctance for hospital attendance and/or admissions thereby missing early warning signs and possible opportunities to mitigate morbidity and potentially mortality.

Amongst the interviewees, there are differing opinions on the role that feelings of insecurity had on mental health. This was not in line with the scoping review, as it was seen to be a considerable detriment to perinatal mental health. This is backed up by a US study’s findings that of the 2740 surveyed pregnant women, 59.2% and 63.7% were stressed about food running out and losing their job or household income respectively (41). These results provide insight into the potential sources of stress during the pandemic specifically. However, post-pandemic these concerns are likely to continue to be relevant, due to the current cost-of-living crisis, in which several countries are experiencing price increases in daily goods and services (79). Therefore, it is essential that these insecurities are queried and addressed in antenatal appointments, to ensure that perinatal mental health isn’t harmed further.

National lockdowns resulted in months of restricted face-to-face interaction, causing the withdrawal of social support that parents often rely on with a newborn (25). In line with the findings of Amelia Ryan and Carol Barber who illustrated the significance of the saying, ‘it takes a village to raise a child’ (37), Participant 4 agreed with this fact by exploring the diverse responsibilities of parenthood, from providing sustenance and comfort to engaging in play, and emphasised the significance of distributing these duties effectively. It is important to highlight this fact as, according to Participant 4, women who experienced childbirth amidst the pandemic are presently expecting their second child, and as previously stated by Participant 5, new parents are accustomed to the isolation of the pandemic and so were unfamiliar with the significance of the support their social network could provide, and so it is vital that they are informed of the value of seeking help from others.

Maternal grief was an outcome of missed pregnancy and postnatal experiences and the relevance of this on maternal mental health was shared by the interviewees as well as the scoping review; the KIIs described this as a “shattering of expectations” that eventually led to depression and anger. This correlates with concepts demonstrated in other themes, namely the dismissive manner of hospitals, the non-existent social networks, the extra sources of anxiety, and the financial situation parents found themselves in.

Raising a baby alone, having been victims to a healthcare system working at full capacity, and the withdrawal of their social network, gave rise to feelings of apprehension in many women’s parenting proficiency. Reliance on primordial instincts were discussed in KIIs for managing this trepidation, along with the importance of mothers placing trust in themselves due to the limited support available to them. This finding ties well with previous studies wherein ‘resilience’ was seen as being a positive contributor to mental health outcomes. One study published in 2022 found that practicing coping and relaxation strategies promoted resilience perinatally (33). Developing a psychoeducational intervention promoting and training mothers to be resilient would be a valuable trait to acquire, especially with the ongoing cost-of-living crisis.

## Data Availability

All relevant data are within the manuscript and its Supporting Information files.

## Acknowledgements

We would like to thank the Brighton and Sussex Medical School Independent Research Project module examiners and markers for their constructive feedback on this project.

## Supplementary data

Supplementary Data 1: PubMed database

Supplementary Checklist 2: CASP Qualitative checklist

Supplementary File 3: Interview guide

Supplementary File 4: Ethics approval forms

Supplementary File 5: Journal formatting guidelines

## References

1. Anderson, M., et al., Exploring social complexities of the COVID-19 pandemic on maternal anxiety: A mixed-methods observational cohort study. European Journal of Midwifery, 2022. 6(October): p. 1–9.

2. Davenport, M.H., et al., Moms are not OK: COVID-19 and maternal mental health. Frontiers in global women’s health, 2020: p. 1.

3. Huschke, S., S. Murphy-Tighe, and M. Barry, Perinatal mental health in Ireland: a scoping review. Midwifery, 2020. 89: p. 102763.

4. Meaney, S., et al., The impact of COVID-19 on pregnant womens’ experiences and perceptions of antenatal maternity care, social support, and stress-reduction strategies. Women and Birth, 2022. 35(3): p. 307–316.

5. Kinser, P.A., et al., Depression, anxiety, resilience, and coping: the experience of pregnant and new mothers during the first few months of the COVID-19 pandemic. Journal of women’s health, 2021. 30(5): p. 654–664.

6. Howard, L.M. and H. Khalifeh, Perinatal mental health: a review of progress and challenges. World Psychiatry, 2020. 19(3): p. 313–327.

7. Bauer, A., M. Knapp, and M. Parsonage, Lifetime costs of perinatal anxiety and depression. Journal of affective disorders, 2016. 192: p. 83–90.

8. Arksey, H. and L. O’Malley, Scoping studies: towards a methodological framework. International journal of social research methodology, 2005. 8(1): p. 19–32.

9. Braun, V. and V. Clarke, Using thematic analysis in psychology. Qualitative research in psychology, 2006. 3(2): p. 77–101.

10. Dib, S., et al., Maternal mental health and coping during the COVID-19 lockdown in the UK: Data from the COVID-19 New Mum Study. International Journal of Gynecology & Obstetrics, 2020. 151(3): p. 407–414.

11. Pope, J., et al., Prenatal stress, health, and health behaviours during the COVID-19 pandemic: An international survey. Women and Birth, 2022. 35(3): p. 272–279.

12. Brown, A. and N. Shenker, Experiences of breastfeeding during COVID-19: Lessons for future practical and emotional support. Maternal & child nutrition, 2021. 17(1): p. e13088.

13. Ceulemans, M., et al., Mental health status of pregnant and breastfeeding women during the COVID-19 pandemic—A multinational cross-sectional study. Acta obstetricia et gynecologica Scandinavica, 2021. 100(7): p. 1219–1229.

14. Gildner, T.E., et al., Associations between postpartum depression and assistance with household tasks and childcare during the COVID-19 pandemic: evidence from American mothers. BMC Pregnancy and Childbirth, 2021. 21(1): p. 1–14.

15. Loret de Mola, C., et al., Maternal mental health before and during the COVID-19 pandemic in the 2019 Rio Grande birth cohort. Brazilian Journal of Psychiatry, 2021. 43: p. 402–406.

16. Masters, G.A., et al., Impact of the COVID-19 pandemic on mental health, access to care, and health disparities in the perinatal period. Journal of psychiatric research, 2021. 137: p. 126–130.

17. Wall, S. and M. Dempsey, The effect of COVID-19 lockdowns on women’s perinatal mental health: a systematic review. Women and Birth, 2022.

18. Barbosa-Leiker, C., et al., Stressors, coping, and resources needed during the COVID-19 pandemic in a sample of perinatal women. BMC pregnancy and childbirth, 2021. 21(1): p. 1–13.

19. Burgess, A., et al., Impact of COVID-19 on pregnancy worry in the United States. Birth, 2022. 49(3): p. 420–429.

20. Liu, C.H., et al., Subjective social status, COVID-19 health worries, and mental health symptoms in perinatal women. SSM-population health, 2022. 18: p. 101116.

21. Ho-Fung, C., et al., Self-reported mental health status of pregnant women in Sweden during the COVID-19 pandemic: a cross-sectional survey. BMC pregnancy and childbirth, 2022. 22(1): p. 1–12.

22. Kara, P., et al., Post-traumatic stress disorder and affecting factors in pregnant women in the COVID-19 pandemic. Psychiatria Danubina, 2021. 33(2): p. 231–239.

23. Kotlar, B., et al., The impact of the COVID-19 pandemic on maternal and perinatal health: a scoping review. Reproductive health, 2021. 18: p. 1–39.

24. Vazquez-Vazquez, A., et al., The impact of the Covid-19 lockdown on the experiences and feeding practices of new mothers in the UK: Preliminary data from the COVID-19 New Mum Study. Appetite, 2021. 156: p. 104985.

25. Jackson, L., et al., Postpartum women’s psychological experiences during the COVID-19 pandemic: a modified recurrent cross-sectional thematic analysis. BMC pregnancy and childbirth, 2021. 21(1): p. 1–16.

26. Anderson, E., et al., Pregnant women’s experiences of social distancing behavioural guidelines during the Covid-19 pandemic ‘lockdown’in the UK, a qualitative interview study. BMC Public Health, 2021. 21: p. 1–12.

27. Zhou, J., et al., Changes in social support of pregnant and postnatal mothers during the COVID-19 pandemic. Midwifery, 2021. 103: p. 103162.

28. Filippetti, M.L., A.D.F. Clarke, and S. Rigato, The mental health crisis of expectant women in the UK: effects of the COVID-19 pandemic on prenatal mental health, antenatal attachment and social support. BMC pregnancy and childbirth, 2022. 22(1): p. 1–10.

29. Jackson, L., et al., Postpartum women’s experiences of social and healthcare professional support during the COVID-19 pandemic: A recurrent cross-sectional thematic analysis. Women and Birth, 2022. 35(5): p. 511–520.

30. Camoni, L., et al., The Impact of the COVID-19 Pandemic on Women’s Perinatal Mental Health: Preliminary Data on the Risk of Perinatal Depression/Anxiety from a National Survey in Italy. International journal of environmental research and public health, 2022. 19(22): p. 14822.

31. Anderson, M.R., et al., Stress, coping and silver linings: How depressed perinatal women experienced the COVID-19 pandemic. Journal of Affective Disorders, 2022. 298: p. 329–336.

32. Bridle, L., et al., Supporting perinatal mental health and wellbeing during COVID19. International journal of environmental research and public health, 2022. 19(3): p. 1777.

33. Thomson, G., et al., Resilience and post-traumatic growth in the transition to motherhood during the COVID-19 pandemic: A qualitative exploratory study. Scandinavian Journal of Caring Sciences, 2022. 36(4): p. 1143–1155.

34. McKinlay, A.R., D. Fancourt, and A. Burton, Factors affecting the mental health of pregnant women using UK maternity services during the COVID-19 pandemic: a qualitative interview study. BMC pregnancy and childbirth, 2022. 22(1): p. 1–15.

35. Fallon, V., et al., Psychosocial experiences of postnatal women during the COVID19 pandemic. A UK-wide study of prevalence rates and risk factors for clinically relevant depression and anxiety. Journal of psychiatric research, 2021. 136: p. 157–166.

36. Koyucu, R.G. and P.P. Karaca, The Covid 19 outbreak: maternal mental health and associated factors. Midwifery, 2021. 99: p. 103013.

37. Ryan, A. and C. Barber, Postnatal depression and anxiety during the COVID-19 pandemic: The needs and experiences of New Zealand mothers and health care providers. Midwifery, 2022. 115: p. 103491.

38. Jones, K., et al., A qualitative analysis of feelings and experiences associated with perinatal distress during the COVID-19 pandemic. BMC Pregnancy and Childbirth, 2022. 22(1): p. 1–19.

39. Lega, I., et al., The psychological impact of COVID-19 among women accessing family care centers during pregnancy and the postnatal period in Italy. International Journal of Environmental Research and Public Health, 2022. 19(4): p. 1983.

40. Yakupova, V., A. Suarez, and A. Kharchenko, Birth Experience, Postpartum PTSD and Depression before and during the Pandemic of COVID-19 in Russia. International journal of environmental research and public health, 2022. 19(1): p. 335.

41. Moyer, C.A., et al., Pregnancy-related anxiety during COVID-19: a nationwide survey of 2740 pregnant women. Archives of women’s mental health, 2020. 23: p. 757–765.

42. Sakalidis, V.S., et al., Longitudinal changes in wellbeing amongst breastfeeding women in Australia and New Zealand during the COVID-19 pandemic. European journal of pediatrics, 2022. 181(10): p. 3753–3766.

43. Brik, M., et al., Psychological impact and social support in pregnant women during lockdown due to SARS-CoV2 pandemic: A cohort study. Acta obstetricia et gynecologica Scandinavica, 2021. 100(6): p. 1026–1033.

44. Javaid, S., et al., The impact of COVID-19 on prenatal care in the United States: Qualitative analysis from a survey of 2519 pregnant women. Midwifery, 2021. 98: p. 102991.

45. Morniroli, D., et al., Exploring the impact of restricted partners’ visiting policies on non-infected mothers’ mental health and breastfeeding rates during the covid-19 pandemic. International Journal of Environmental Research and Public Health, 2021. 18(12): p. 6347.

46. Zhang, C.X.W., et al., Evaluating depression and anxiety throughout pregnancy and after birth: impact of the COVID-19 pandemic. American Journal of Obstetrics & Gynecology MFM, 2022. 4(3): p. 100605.

47. Chapman, G.E., I. Ishlek, and J. Spoors, Google search behaviour relating to perinatal mental wellbeing during the United Kingdom’s first COVID-19 lockdown period: a warning for future restrictions. Archives of Women’s Mental Health, 2021. 24: p. 681–686.

48. Perez, A., et al., Experience of early motherhood during the first wave of the COVID-19 pandemic in Northern Germany: a single-centre before and after comparison. Journal of reproductive and infant psychology, 2021: p. 1–17.

49. Jeličić, L., et al., Maternal Anxiety and Its Associated Factors During the First and Second Wave of COVID-19 Pandemic in Serbia: A Cross-Sectional Study. Psychology Research and Behavior Management, 2022: p. 3775–3792.

50. Lequertier, B., et al., Perinatal depression in Australian women during the COVID19 pandemic: The birth in the time of COVID-19 (BITTOC) study. International journal of environmental research and public health, 2022. 19(9): p. 5062.

51. Lin, H.-C., et al., Maternal self-efficacy buffers the effects of COVID-19–related experiences on postpartum parenting stress. Journal of Obstetric, Gynecologic & Neonatal Nursing, 2022. 51(2): p. 177–194.

52. Farewell, C.V., et al., A mixed-methods pilot study of perinatal risk and resilience during COVID-19. Journal of Primary Care & Community Health, 2020. 11: p. 2150132720944074.

53. Corno, G., et al., The role of perceived social support on pregnant women’s mental health during the COVID-19 pandemic. Journal of Reproductive and Infant Psychology, 2022: p. 1–15.

54. Orsolini, L., et al., Fear and anxiety related to COVID-19 pandemic may predispose to perinatal depression in Italy. Frontiers in psychiatry, 2022. 13.

55. Groulx, T., et al., Prenatal care disruptions and associations with maternal mental health during the COVID-19 pandemic. Frontiers in Global Women’s Health, 2021. 2: p. 648428.

56. Grumi, S., et al., Depression and anxiety in mothers who were pregnant during the COVID-19 outbreak in Northern Italy: the role of pandemic-related emotional stress and perceived social support. Frontiers in Psychiatry, 2021. 12: p. 716488.

57. Pacheco, F., et al., Breastfeeding during COVID-19: a narrative review of the psychological impact on mothers. Behavioral Sciences, 2021. 11(3): p. 34.

58. Preis, H., et al., Vulnerability and resilience to pandemic-related stress among US women pregnant at the start of the COVID-19 pandemic. Social science & medicine, 2020. 266: p. 113348.

59. Larotonda, A. and K.A. Mason, New life, new feelings of loss: journaling new motherhood during Covid-19. SSM-Mental Health, 2022. 2: p. 100120.

60. Doyle, F.L. and L. Klein, Postnatal depression risk factors: an overview of reviews to inform COVID-19 research, clinical, and policy priorities. Frontiers in Global Women’s Health, 2020. 1: p. 577273.

61. Goldstein, E., et al., Latent class analysis of health, social, and behavioral profiles associated with psychological distress among pregnant and postpartum women during the COVID-19 pandemic in the United States. Birth, 2022.

62. Hildersley, R., et al., Changes in the identification and management of mental health and domestic abuse among pregnant women during the COVID-19 lockdown: regression discontinuity study. BJPsych open, 2022. 8(4): p. e96.

63. Liu, C.H., et al., Unexpected changes in birth experiences during the COVID-19 pandemic: Implications for maternal mental health. Archives of gynecology and obstetrics, 2021: p. 1–11.

64. Shuman, C.J., et al., “Mourning the experience of what should have been”: experiences of peripartum women during the COVID-19 pandemic. Maternal and child health journal, 2022: p. 1–8.

65. Hendrix, C.L., et al., Geotemporal analysis of perinatal care changes and maternal mental health: an example from the COVID-19 pandemic. Archives of Women’s Mental Health, 2022. 25(5): p. 943–956.

66. Zhang, C.J.P., et al., Psychobehavioral responses, post-traumatic stress and depression in pregnancy during the early phase of COVID-19 Outbreak. Psychiatric research and clinical practice, 2021. 3(1): p. 46–54.

67. Zhang, Y. and Z.F. Ma, Psychological responses and lifestyle changes among pregnant women with respect to the early stages of COVID-19 pandemic. International Journal of Social Psychiatry, 2021. 67(4): p. 344–350.

68. Wolfe-Sherrie, E.J., et al., “Hey child, why were you born when the world is almost over?”: An analysis of first-time mothers’ postpartum experiences during the early stages of the COVID-19 pandemic in Coatepec, Veracruz, Mexico. Maternal and Child Health Journal, 2022. 26(8): p. 1732–1740.

69. An, R., et al., A survey of postpartum depression and health care needs among Chinese postpartum women during the pandemic of COVID-19. Archives of psychiatric nursing, 2021. 35(2): p. 172–177.

70. Motrico Martínez, E., et al., The impact of the COVID-19 pandemic on perinatal depression and anxiety: a large cross-sectional study in Spain. Psicothema, 2022.

71. Mayhew, K. and P. Anand, COVID-19 and the UK labour market. Oxford Review of Economic Policy, 2020. 36(Supplement_1): p. S215–S224.

72. Chen, Q., et al., Prevalence and risk factors associated with postpartum depression during the COVID-19 pandemic: a literature review and meta-analysis. International journal of environmental research and public health, 2022. 19(4): p. 2219.

73. Wilson, C.A., et al., Challenges and opportunities of the COVID-19 pandemic for perinatal mental health care: a mixed-methods study of mental health care staff. Archives of Women’s Mental Health, 2021. 24(5): p. 749–757.

74. Ahmad, M. and L. Vismara, The psychological impact of COVID-19 pandemic on women’s mental health during pregnancy: A rapid evidence review. International Journal of Environmental Research and Public Health, 2021. 18(13): p. 7112.

75. Lobel, M., et al., Common model of stress, anxiety, and depressive symptoms in pregnant women from seven high-income Western countries at the COVID-19 pandemic onset. Social Science & Medicine, 2022. 315: p. 115499.

76. Kakaraparthi, V.N., et al., Anxiety, depression, worry, and stress-related perceptions among antenatal women during the COVID-19 pandemic: Single group repeated measures design. Indian Journal of Psychiatry, 2022. 64(1): p. 64.

77. Xu, K., et al., Mental health among pregnant women under public health interventions during COVID-19 outbreak in Wuhan, China. Psychiatry research, 2021. 301: p. 113977.

78. Liu, C.H., C. Erdei, and L. Mittal, Risk factors for depression, anxiety, and PTSD symptoms in perinatal women during the COVID-19 Pandemic. Psychiatry research, 2021. 295: p. 113552.

79. Andersen, K. and A. Reeves, The cost of living crisis is harming mental health, partly because of previous cuts to social security. 2022, British Medical Journal Publishing Group.

80. Pannucci, C.J. and E.G. Wilkins, Identifying and avoiding bias in research. Plastic and reconstructive surgery, 2010. 126(2): p. 619.

81. Drucker, A.M., P. Fleming, and A.-W. Chan, Research techniques made simple: assessing risk of bias in systematic reviews. Journal of Investigative Dermatology, 2016. 136(11): p. e109–e114.

